# Coronavirus epidemic: prediction and controlling measures

**DOI:** 10.1101/2020.04.11.20062125

**Authors:** Mohammad Mehdi Pejman, Armaghan Fereidooni

## Abstract

The COVID-19 outbreak has caused over 1.7 million (still increasing) confirmed cases globally as of April 10th, 2020. The levels of spread and severity of the virus lead to a wide-spread political and economic turmoil. We believe that two critical contributing factors need to be taken into account by the authorities to make effective decisions for controlling the spread of the virus: (*i*) being familiar with the most effective controlling measures and (*ii*) having a mathematical model to predict the spread of the virus. In this study, we provided information regarding both of these crucial factors. First, we investigated the importance of different measures such as quarantine, isolation, face mask, social distancing, etc. in controlling the virus in various countries. We then present a mathematical model to predict the spread of the virus in different countries. Our prediction shows an excellent match with the actual data up to now.

## 1. Introduction

At the end of 2019, the COVID-19 outbreak originated from Wuhan, a city in central China. The outbreak leads to confinement of over 1 billion people to their homes since the end of January 2020 and indeed tremendously disrupts the health care, economy, and well being. After a while, as the circumstances in China appears to be stabilizing, dramatic rises in confirmed cases are being reported in South Korea, Italy, Japan, and Iran. The spread of the virus continues worldwide, and eventually, on March, 11th 2020, the world health organization (WHO) characterized the 2019 outbreak of coronavirus disease (COVID-19) as a pandemic [1]. According to the World Health Organization (WHO), as of April, 10th 2020, there were around 1.7 million confirmed cases and around 103 thousand deaths from more than 140 countries.

The unprecedented levels of spread and severity lead to creating a wide-spread political and economic turmoil. The main reason of the great amount of disturbance, confusion, and uncertainty regarding this virus backs to the fact that the majority of infected people do not experience severe symptoms ^1^, as a result making it more likely for them to remain mobile, and hence to infect others. COVID-19 is shown to have a globally averaged fatality ratio of about 6.06% to date. In comparison, in late 2002 and early 2003, a similar outbreak happened with the occurrence of a severe acute respiratory syndrome (SARS). Although the etiological agent of SARS is also a coronavirus, the virus was not able to spread as widely as in the COVID-19. The possible reason for the SARS outbreak to be less devastating compares to the COVID-19 is, paradoxically, because of its much higher fatality ratio (around 10% globally), making it too severe to spread readily. Thus, it is probable that this specific combination of traits has made the COVID-19 outbreak one of the largest in history. Generally, the total number of deaths from the pneumonia-related disease is low and the majority of them had health conditions such as hypertension, diabetes, or cardiovascular disease that weakened their immune systems.

In comparison with the seventeen years ago, when the world had to confront the SARS pandemics, substantial progress has been achieved. The scientists in the Chinese Center for Disease Control and Prevention (CDC) networks identified a new strain of coronavirus and recognized it as the pathogen of this novel epidemic within a few weeks after the first case was noticed. The Chinese health authorities communicated the detail information of the epidemic with the WHO and the international community and shared the full genome sequence of the novel virus with the global scientific community. Despite the substantial progress being made, there are still a lot of issues remain to deal with, and significant challenges have been imposed to the public health emergency response globally. The potential of the health care systems in different countries such as the number of available ICU beds, ventilators, test kits, masks, etc can be deemed as crucial factors in controlling the spread and mortality rates of the COVID-19 globally. On the other hand, trips and traditional events in various countries are the main contributing factors in increasing the spread rate of COVID-19. Thus, it is not deniable that controlling the epidemic requires a coordinated and integrated international response. As recognized by WHO, mathematical models, especially those timely, play a key role in informing evidence-based decisions by health decision and policy makers.

Many different mathematical models are presented in the literature to predict the spread of COVID-19 such as SIRD approach [5], SEIR method [6], and SIR scheme [7–9]. In this study, we have also used the SIR method to predict the spread of COVID-19 in different countries. We have also investigated the essential protective measures to control the outbreak. Indeed, we discussed the positive and negative aspects of the measures taken by different countries in controlling COVID-19.

The remainder of the paper is organized as follows. In Section 2, we present the mathematical model to predict the pandemic. Section 3 discusses the crucial measures that need to be taken into account by the authorities and the positive and negative aspects of them. Finally, in Section 4, the results of our mathematical model prediction are presented for several countries.

## 2. Mathematical model

In this study, we use one of the most classic epidemic models, namely the SIR model. Three states are defined in this model:

- S(t): the number of individuals susceptible of contracting the infection at time t;
- I(t): the number of active infected individuals at time t;
- R(t): the number of individuals who are recovered at time t.

The main governing equations of this model are as follows:

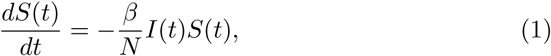

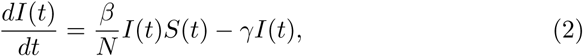

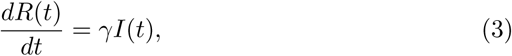

where t is time, N is the total population size (*N* = *S*(*t*)+ *I*(*t*)+ *R*(*t*) = *const*.), *β* ∈ {ℝ > 0} is the infection rate, and *γ* ∈ {ℝ > 0} is the recovery rate.The initial conditions of the governing equations (1-3) are *S*(0) = *S*_0_ > 0, *I*(0) = *I*_0_ ≥ 0, and *R*(0) = 0, respectively.

Using the following minimization problem, we can find the unknown parameters *β*, and *γ* to fit the actual data.

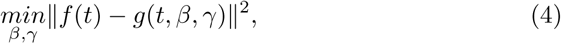

where *β* and *γ* are design parameters, *f*(*t*) = *I*_*actual*_ + *R*_*actual*_ is the summation of cumulative numbers of the actual number of infected people and recovered ones up to time t, and *g*(*t, β, γ*) = *I*(*t*) + *R*(*t*) is the summation of the predicted values for cumulative numbers of infected people and recovered ones until time t. Note that the design parameters of this minimization problem are the unknowns *β* and *γ*.

Now that we have the values of *β* and *γ*, we can use the SIR model to predict the COVID-19 spread behavior in the future. To do so, we introduce additional equations with respect to the expected behavior when the time approaches infinity (*t → ∞;*). We know that as the time approaches infinity, *I*(*∞;*) = 0, thus *R*(*∞;*) can be given by

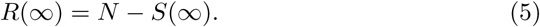

Using (3) and (1), *S*(*∞;*) is

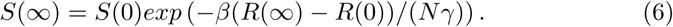

By plugging in (6) in (5), we obtain *R*(*∞;*) as

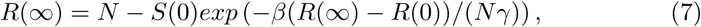

which is a nonlinear equation. By solving this nonlinear equation, we can readily predict the behavior of S(t), I(t), and R(t)

## 3. Importance of protective measures

Figure 1(a) shows the number of confirmed cases for the eight countries of China, Germany, Iran, Italy, South Korea, Spain, the UK, and the US until April, 10th, 2020 based on the data reported in [10]. The horizontal axis in this figure is the days that the first confirmed case is found in each one of these countries. Of course, the number of confirmed cases is a function of the number of tests which have been done by each country, the countries’ population, etc. Obviously, there is significant uncertainty and underestimation in the actual number of individuals who currently have the disease. Keep in mind that there are also many individuals who do not show severe symptoms when they get the disease. Thus, the number of infected people is not an appropriate measure to determine how much the virus spreads in each country.

**Figure 1:**
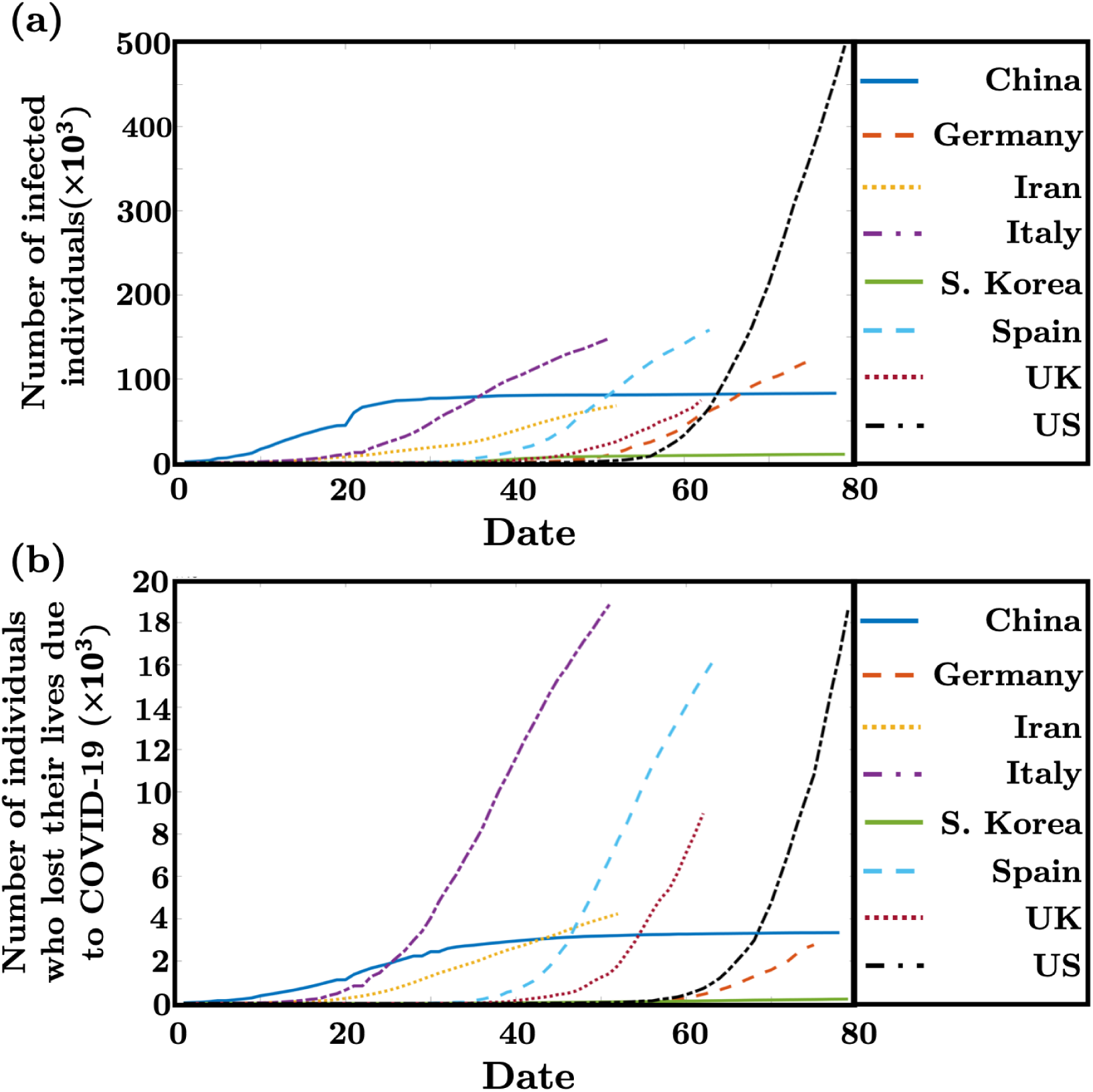
(a) Number of confirmed cases and (b) number of individuals who lost their lives due to COVID-19 for the eight countries of China, Germany, Iran, Italy, South Korea, Spain, the UK, and the US until April, 10th, 2020 based on the data reported in [10].

We believe that the number of individuals who lost their lives due to COVID-19 can be count as a more trustable measure with a lower uncertainty in comparison with the number of confirmed cases to compare the spread of the virus in different countries with each other. Thus, in Fig. 1(b), we present the number of individuals who lost their lives because of COVID-19 in different countries. The results reveal that there is a clear difference in the slope of the curves in China, South Korea with the other countries. This behavior can be correlated to the protective measures that these countries take into account to control the spread of COVID-19. Since at this time there are no confirmed treatments for this infection, prevention is crucial. Thus, it is of great importance to go through the protective measures that the countries can perform to reduce the spread rate of the virus.

COVID-19 has several features which make the prevention process quite difficult such as the infectivity even before the start of symptoms in the incubation period, relatively long incubation period, transmission from asymptomatic indi-viduals, tropism for mucosal surfaces, long duration of the illness, and also the possibility of transmission even after recovery [1, 11, 12].

The first prevention measure which is quite effective to be performed in the very early stages of the spreading of the virus in a country is isolation and quarantine ^2^. Quarantine has been always one of the measures for the outbreaks such as the 14th-century Italian “quarantino” to control the plague (Black Death), SARS outbreak, and etc. In the case of COVID-19, as it is reported in [14], the median incubation period is 7-day for adults based on the data obtained from 2015 laboratory-confirmed COVID-19 cases including 99 children in 28 Chinese provinces. They suggested that the extension of the adult quarantine period to 18 days or 21 days could be more effective in preventing virus-spreading and controlling the disease. Even though effective in controlling the spread rate of the virus, quarantine may result in adverse psychological issues. For instance, it is reported that some people quarantined during the SARS outbreak experienced symptoms of post-traumatic stress disorder and depression [15, 16]. Moreover, people may experience financial hardship. Thus, it is critical for the authorities to make sure quarantined people have ongoing access to resource materials, financial aids, and open lines of communication and psychosocial support [15, 17]. For example, in the US, the Senate on March, 25th, 2020 approved a historic, $2 trillion stimulus package to provide a jolt to an economy reeling from the coronavirus pandemic which can be considered as an effective measure to alleviate the financial difficulties of the people during the quarantine.

The other important measure is maintaining physical distancing. For example, in Wuhan, China, they enforced the physical distancing measures, including but not limited to school and workplace closures and health promotions that en-courage the general public to avoid crowded places. Moreover, performing a lot of tests to recognize the infected individuals can be really helpful in controlling the spread of the virus. South Korea did a pretty well job in this matter.

One of the measures which have been a matter of controversy among different countries and organizations is the essence of using face masks. The countries have various recommendations on face mask use in community settings. Since the outbreak of SARS, the use of face masks has become ubiquitous in China, South Korea, Japan, Hong Kong, Singapore, and some other Asian countries. Some provinces in China have enforced compulsory face mask policies in public; however, China’s national guideline has adopted a risk-based approach in offering recommendations for using face masks. There is a consistency in the recommendation that symptomatic people and those in health-care settings should use face masks, but discrepancies are observed in the community settings. We believe one of the main reasons that some of the countries would not recommend the entire population of the country to use face masks is the shortage in the number of available masks. They would give priority to healthcare workers. For example, the US Surgeon General advised against buying and using masks by healthy people [18]. It makes sense to some extent since the highest risk in COVID-19 is transmission to healthcare workers. In the SARS outbreak of 2002, 21% of the infected ones were healthcare workers [19]. To the best of our knowledge, there is not any clear evidence that face masks can provide effective protection against respiratory infections in the community. However, there is evidence that COVID-19 has the potential to be transmitted before the start of the symptom. Thus, community transmission may be decreased if everyone wears face masks. It is quite interesting that in countries such as Japan, South Korea, Hong Kong, and Singapore where wearing the face masks is enforced, the spread of the COVID-19 is controlled pretty well. For sure, using the face masks is the only reason for the great control seen in these countries but it can be deemed as one of the effective measures. Noteworthy that improper use of face masks, such as not altering disposable masks, has the potential to jeopardize the protective effect and even lead to increasing the risk of infection. It is essential to rise the public knowledge regarding how to use the mask and how to dispose of it to reduce the risk of infection from the face mask itself.

Noteworthy that individuals who have hypertension, diabetes, COPD, cardiovascular disease, and cerebrovascular disease are at a higher risk. Authorities need to educate the public about the risk of getting COVID-19 in these patients. Furthermore, educating the public in terms of practicing social distancing, washing hands regularly, avoiding touching the face, avoiding travel, etc. can be quite helpful in controlling the spread of the virus.

## 4. Prediction

We applied the proposed mathematical model to the data provided by [10] to predict the spread of the COVID-19 in the upcoming days. The prediction of total and the daily number of infected individuals by the SIR model for South Korea, Spain, the UK, the US, China, Germany, Iran, and Italy are shown in Fig. 2 and 3. The results show that countries such as China, Germany, Italy, South Korea, and Spain are quite close to plateau their curve. Of course, there are many sources of uncertainty in the data and every mathematical model highly depends on the accuracy of the input data. But in general, the proposed mathematical model shows a promising potential in predicting the outbreak. Up to now (April, 10th, 2020), our prediction shows an excellent match with the actual data.

**Figure 2:**
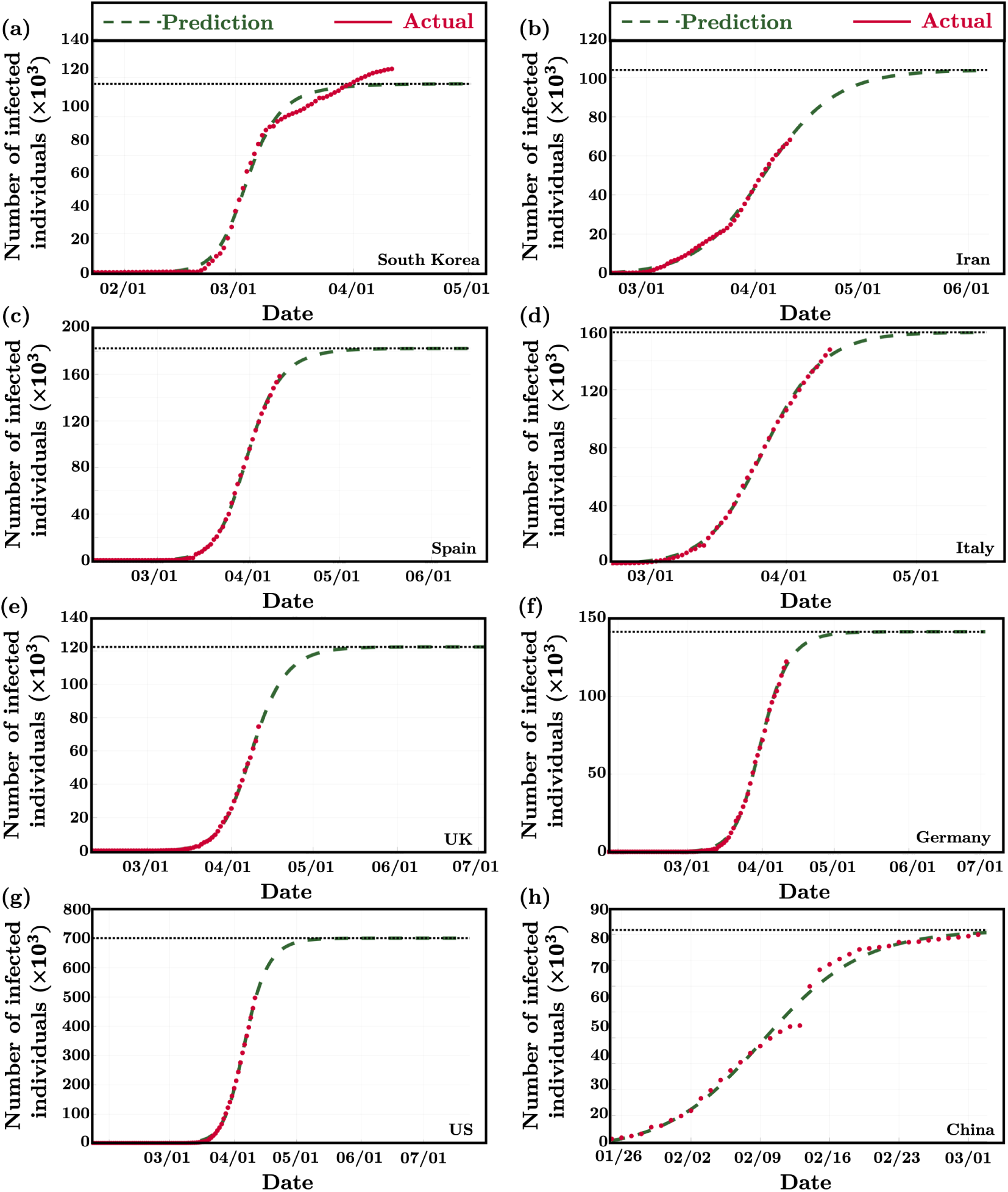
Prediction of the total number of confirmed cases for (a) South Korea, (b) Iran, (c) Spain, (d) Italy, (e) UK, (f) Germany, (g) US, and (h) China. The results are obtained based on the data reported in [10] until April, 10th, 2020.

**Figure 3:**
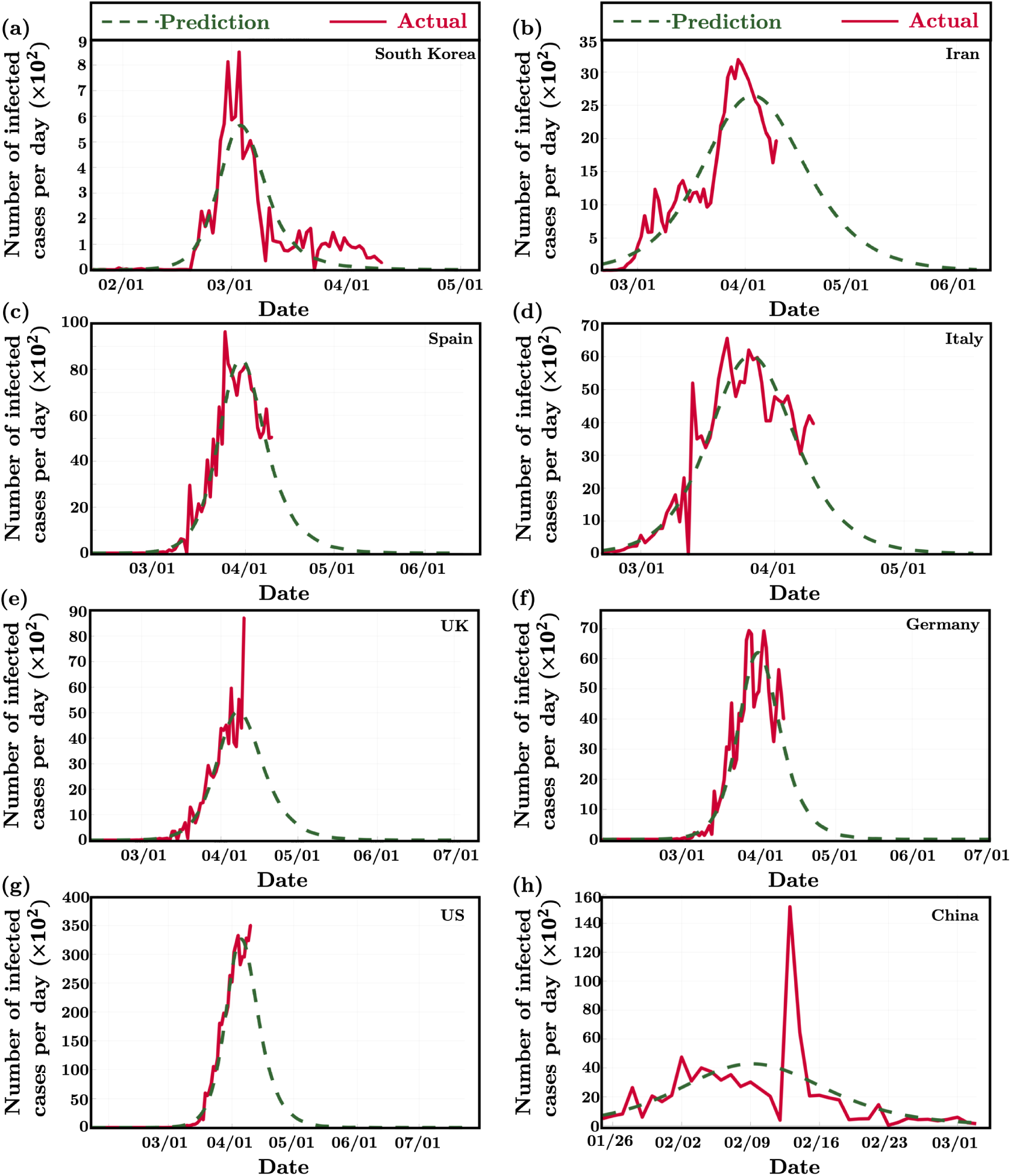
Prediction of the daily number of confirmed cases for (a) South Korea, (b) Iran, (c) Spain, (d) Italy, (e) UK, (f) Germany, (g) US, and (h) China. The results are obtained based on the data reported in [10] until April, 10th, 2020. The results are obtained based on the data reported in [10] until April, 10th, 2020.

It is worth noting that for the countries that the slope of the curve is getting negative, it is quite important to maintain the measures that they considered to control the outbreak. This is due to the fact that the second peak might happen in the situation that the authorities in these countries would not maintain the controlling measures anymore. To prevent the occurrence of the second peak especially in the countries that enforced quarantine measures, it is crucial to release the quarantine order in an appropriate time. They should consider the slope of the increase in the number of daily confirmed cases, the potential of the health-care system, and the number of daily screening tests.

## 5. Conclusion

COVID-19 can be count as an extraordinary challenge globally. Each decision makes by the authorities all around the world can affect the controlling procedure of the virus. Thus, it is of great importance to investigate on the critical and effective contributing factors in controlling the outbreak, and also forecasting the spread of the virus in the near future. In this study, we provided information regarding both of these contributing factors. We discussed the importance of different measures such as quarantine, isolation, face mask, social distancing, etc. in controlling the spread of the virus in various countries. We stress that educating the public in terms of practicing social distancing, washing hands regularly, avoiding touching the face, avoiding travel, etc. can be quite helpful in controlling the spread of the virus. Finally, We present a mathematical model to predict the spread of the virus in different countries. Our prediction shows an excellent match with the actual data up to now.

## Data Availability

All data are available per request.

## Footnotes

1 The common symptoms of COVID-19 are fever, cough, fatigue, sputum production, and headache [2–4].

2 Isolation is typically used for infected person and quarantine is for the person who is suspected to have the disease [13].

## References

1. World health organization. coronavirus disease [covid-19] technical guidance: Infection prevention and control. available at: https://www.who.int/emergencies/diseases/novel-coronavirus-2019/technical-guidance/infection-prevention-and-control.

2. K. Ma, T. Chen, M. Han, W. Guo, Q. Ning, Management and clinical thinking of coronavirus disease 2019, Zhonghua gan zang bing za zhi= Zhonghua ganzangbing zazhi= Chinese journal of hepatology 28 (2020) E002.

3. Z. Zhu, C. Zhong, K. Zhang, C. Dong, H. Peng, T. Xu, A. Wang, Z. Guo, Y. Zhang, Epidemic trend of corona virus disease 2019 (covid-19) in mainland china, Zhonghua yu fang yi xue za zhi [Chinese journal of preventive medicine] 54 (2020) E022–E022.

4. H. Yang, G. Duan, Analysis on the epidemic factors for the corona virus disease, Zhonghua yu Fang yi xue za zhi [Chinese Journal of Preventive Medicine] 54 (2020) E021–E021.

5. C. Anastassopoulou, L. Russo, A. Tsakris, C. Siettos, Data-based analysis, modelling and forecasting of the covid-19 outbreak, PloS one 15 (3) (2020) e0230405.

6. C. Zhan, C. Tse, Y. Fu, Z. Lai, H. Zhang, Modelling and prediction of the 2019 coronavirus disease spreading in china incorporating human migration data, Available at SSRN 3546051.

7. F. Zullo, Some numerical observations about the covid-19 epidemic in italy, arXiv preprint 2003.11363.

8. I. Nesteruk, Estimations of the coronavirus epidemic dynamics in south korea with the use of sir model, Preprint.] ResearchGate.

9. M. Batista, Estimation of the final size of the covid-19 epidemic, Preprint.] medRxiv.

10. https://coronavirus.jhu.edu/map.html.

11. Y.-H. Jin, L. Cai, Z.-S. Cheng, H. Cheng, T. Deng, Y.-P. Fan, C. Fang, D. Huang, L.-Q. Huang, Q. Huang, et al., A rapid advice guideline for the diagnosis and treatment of 2019 novel coronavirus (2019-ncov) infected pneumonia (standard version), Military Medical Research 7 (1) (2020) 4.

12. T. Singhal, A review of coronavirus disease-2019 (covid-19), The Indian Journal of Pediatrics (2020) 1–6.

13. E. A. Coomes, J. A. Leis, W. L. Gold, Quarantine, CMAJ 192 (13) (2020) E338–E338.

14. X. Jiang, Y. Niu, X. Li, L. Li, W. Cai, Y. Chen, B. Liao, E. Wang, Is a 14-day quarantine period optimal for effectively controlling coronavirus disease 2019 (covid-19)?, medRxiv.

15. S. K. Brooks, R. K. Webster, L. E. Smith, L. Woodland, S. Wessely, N. Greenberg, G. J. Rubin, The psychological impact of quarantine and how to reduce it: rapid review of the evidence, The Lancet.

16. T. Day, A. Park, N. Madras, A. Gumel, J. Wu, When is quarantine a useful control strategy for emerging infectious diseases?, American Journal of Epidemiology 163 (5) (2006) 479–485.

17. L. Hawryluck, W. L. Gold, S. Robinson, S. Pogorski, S. Galea, R. Styra, Sars control and psychological effects of quarantine, toronto, canada, Emerging Infectious Diseases 10 (7) (2004) 1206.

18. S. Feng, C. Shen, N. Xia, W. Song, M. Fan, B. J. Cowling, Rational use of face masks in the covid-19 pandemic, The Lancet Respiratory Medicine.

19. D. Chang, H. Xu, A. Rebaza, L. Sharma, C. S. D. Cruz, Protecting health-care workers from subclinical coronavirus infection, The Lancet Respiratory Medicine 8 (3) (2020) e13.

